# Genetic variation in HLA, IGKV and HHEX loci influence mRNA vaccine-induced long-lasting humoral protection against COVID-19

**DOI:** 10.1101/2025.10.21.25337971

**Authors:** Barbara Puzek, Paul R. Wratil, Christian Janke, Thu Giang Le Thi, Raquel Rubio-Acero, Gaia Lupoli, Irene Charlotte Schoof, Marcel Stern, Ana Zhelyazkova, Jochen Rech, Alexander Choukér, Berthold Koletzko, Andreas Wieser, Helga P. Török, Noemi Castelletti, Oliver T. Keppler, Christof Geldmacher, Veit Hornung, Eleftheria Zeggini, Kristina Adorjan, Michael Hoelscher, Sibylle Koletzko, Sarah Kim-Hellmuth

## Abstract

There is a notable variability in the humoral response mounted across individuals after COVID-19 vaccination. A growing number of studies link genetic factors to antigen-specific antibody levels after the first two immunizations. However, knowledge on the potential influence of genetics on the neutralizing activity of vaccine-induced antibodies and the humoral immune response to the third immunization is limited. Here, we performed genome-wide association studies on antigen-specific antibody concentrations and live-virus neutralization activities after two and three doses of COVID-19 vaccines across two German cohorts of SARS-CoV-2 infection-naïve individuals (RisCoin study n=2877; KoCo19 study n=1654). We found the Human leukocyte antigen (HLA) locus to be associated with differential live-virus neutralizing and anti-SARS-CoV-2 spike antibodies after both the second and third vaccinations. Consistent with its effect on third-dose neutralizing antibodies the HLA locus was further associated with a lower incidence of breakthrough infections, indicating sustained protection. We confirmed the association of the Immunoglobulin kappa variable cluster (IGKV) with antibody concentrations after two vaccinations and discovered a novel association in the Hematopoietically expressed homeobox (HHEX) locus. Studying HLA, IGKV, and HHEX may clarify mechanisms of variable humoral immunity and contribute towards personalized vaccination strategies.

## Introduction

In response to the coronavirus disease 2019 (COVID-19) caused by the emergence of severe acute respiratory syndrome coronavirus 2 (SARS–CoV–2), effective vaccines were developed and rolled out swiftly. After immunization with these COVID-19 vaccines, both cellular and humoral immune responses are evoked.^1–5^ The latter consists of anti-SARS-CoV-2 spike binding and virus neutralizing antibodies. Vaccine-induced humoral immunity to SARS-CoV-2 is largely dependent on virus neutralizing antibodies, the titres of which were shown to correlate with protection from symptomatic and, especially, severe disease.^6^ Among COVID-19-vaccinated individuals, there is, however, a notable variability in the levels and quality of these antibodies^7–11^, affecting their susceptibility to infection and their likelihood of experiencing severe disease. This variation was attributed to a range of factors, including environmental influences and genetic differences.^12–23^ Obtaining a more profound understanding of the inter-individual variability of the humoral immune response after COVID-19 vaccination is key to comprehend and predict immunity to COVID-19.

There is a growing number of studies pointing to genetic effects that influence vaccination-induced anti-SARS-CoV-2 spike antibody response.^14–23^ Multiple studies report associations in the Human Leukocyte Antigen (HLA) class I and II loci, with specific alleles reported differing across studies.^14–23^ Previous analyses focused on genetic determinants solely influencing anti-spike binding antibodies (antibody concentration or binary serostatus). There is, nevertheless, a lack of research on genetic factors influencing antibody-mediated live-virus neutralizing capacities induced by COVID-19 vaccination, which are, indeed, a more accurate correlate of protection from a SARS-CoV-2 infection and severe disease.^6,24,25^ Furthermore, genetic data on humoral immune responses after COVID-19 vaccination was, thus far, limited to the effects of two immunizations. Nowadays, a large proportion of the population has, however, received at least three COVID-19 vaccinations with significant impact on the longevity and the breadth of the induced immune response.^10,26,27^

Here, we performed genome-wide association studies on humoral immune responses in SARS-CoV-2 infection-naïve individuals from two German cohorts that were longitudinally sampled after the second and third COVID-19 vaccinations. Genetic associations not only to anti-spike antibody concentrations but also to live-virus neutralization activities were examined. These analyses revealed that variants at the HLA, IGKV and HHEX loci influence durable immunity against COVID-19. By addressing underexplored aspects of protective and long-lasting immunity and identifying a novel association signal in the HHEX locus, our findings add to the understanding of genetic contributions to the observed variability in COVID-19 vaccine responses. Our research might, thus, be important to identify vulnerable individuals as well as adapt and personalize future COVID-19 and broader mRNA vaccination campaigns.

## Results

### Cohort characteristics

To explore genetic contributions to antibody level variability after two COVID-19 vaccinations, we used data obtained from two cohorts of adult, SARS-CoV-2 infection-naïve COVID-19 vaccinees from Germany, one consisting of healthcare workers (RisCoin)^28,29^ and the other from individuals recruited from the general population of Munich, Bavaria (KoCo19)^30,31^ (**Fig. 1a**). In the sera obtained from both cohorts we measured SARS-CoV-2-specific anti-spike as well as anti-nucleocapsid antibody concentrations, and in the specimens from the RisCoin cohort we, additionally, assessed live-virus neutralization activity against virus variant omicron BA.1. Only those participants were included into the data evaluation who serologically and anamnestically had no prior SARS-CoV-2 infection at the time of sampling. After applying stringent criteria for our data evaluation (**see Methods**), the first cohort (RisCoin) included 2877 individuals who were sampled at a median of 8 months after receiving the second and 1225 participants 2 months after the third vaccination with mRNA vaccines. The second cohort (KoCo19) consisted of 1654 individuals sampled 2 months after the second COVID-19 vaccination. The cohort characteristics are summarized in **Supplementary Table 1**.

**Fig. 1.**
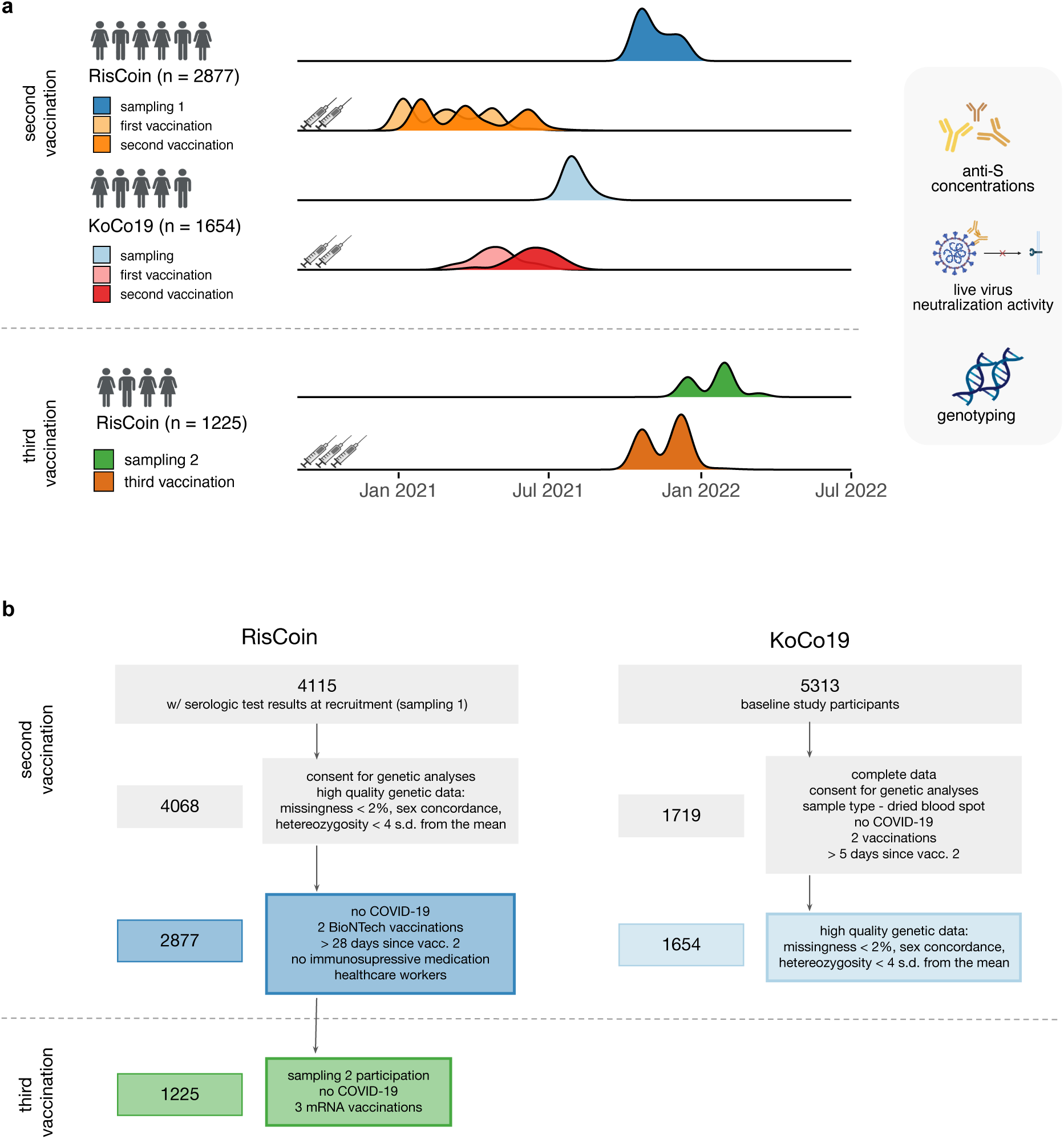
Study timeline and individual selection. **a** Study overview. After the second COVID-19 vaccination, analyses included 2877 individuals from the RisCoin study and 1654 individuals from the KoCo19 study. COVID-19 vaccinations were administered from January to July 2021 and subsequent blood sampling was performed from August 2021 to February 2022. Samples were used to determine anti-SARS-CoV-2 spike antibody concentrations and – in case of the RisCoin cohort – live virus neutralization activities as well as to obtain genotype data. After the third COVID-19 vaccination, analyses included 1225 individuals from the RisCoin study. Blood sampling was performed from December 2021 to May 2022. **b** Flow diagram of participant selection for genetic analyses in RisCoin and KoCo19 study.

### Variation in HLA, IGKV and HHEX loci associated with anti-S antibody concentrations after two COVID-19 vaccinations

To identify possible genetic factors associated with altered humoral immune responses after two COVID-19 vaccinations, we first ran genome-wide association studies (GWAS) on both studies individually. These analyses were performed with inverse-normal transformed antibody level phenotypes and included age, gender, days since the second vaccination, the first five principal components of genetic data and the genetic relatedness matrix as covariates. In both of the individual analyses, we found a genome-wide significant signal (p-value < 5×10^-8^), in the Human Leukocyte Antigen (HLA) region (**Supplementary Fig. 1**, **Table 1**). Next, we performed a meta-analysis of GWAS by combining the two individual studies. In this meta-analysis, we identified four independent genome-wide significant signals across three chromosomes (**Fig. 2a**) and observed no p-value inflation (λ = 1, **Fig. 2b**, inset). The results included three previously reported signals within the HLA and Immunoglobulin kappa variable cluster (IGKV) regions, and a novel genetic signal on chromosome 10 (**Table 1**). The closest gene to the lead variant of the signal on chromosome 10 is Hematopoietically expressed homeobox (HHEX) (**Fig. 2b**), a transcription factor involved in a plethora of biological processes including development and hematopoiesis.^32^

**Table 1.** Summary statistics of the lead variants in four independent loci identified in the GWAS meta-analysis of anti-spike antibody concentrations after two COVID-19 vaccinations. Genetic effect estimates and association p-values are shown per cohort and for the meta-analysis. Position is given with respect to hg19/GRCH37 genomic build. EAF = effect allele frequency.

| Variant | Chromosome | Position | Effect allele | EAF | KoCo19 beta | RisCoin beta | Meta-analysis beta | KoCo19 p-value | RisCoin p-value | Meta-analysis p-value |
| --- | --- | --- | --- | --- | --- | --- | --- | --- | --- | --- |
| rs111357016 | 2 | 89281528 | A | 0.067 | 0.390 | 0.31 | 0.35 | 4.3e-06 | 8.3e-05 | 1.7e-09 |
| rs1655904 | 6 | 29917146 | C | 0.810 | -0.230 | -0.28 | -0.26 | 1.1e-08 | 4.4e-13 | 2.9e-20 |
| rs9271182 | 6 | 32578230 | G | 0.690 | 0.120 | 0.20 | 0.16 | 1.0e-03 | 8.3e-11 | 1.5e-12 |
| rs1544210 | 10 | 94487801 | A | 0.490 | 0.074 | 0.12 | 0.10 | 1.7e-02 | 2.9e-07 | 3.1e-08 |

**Fig. 2.**
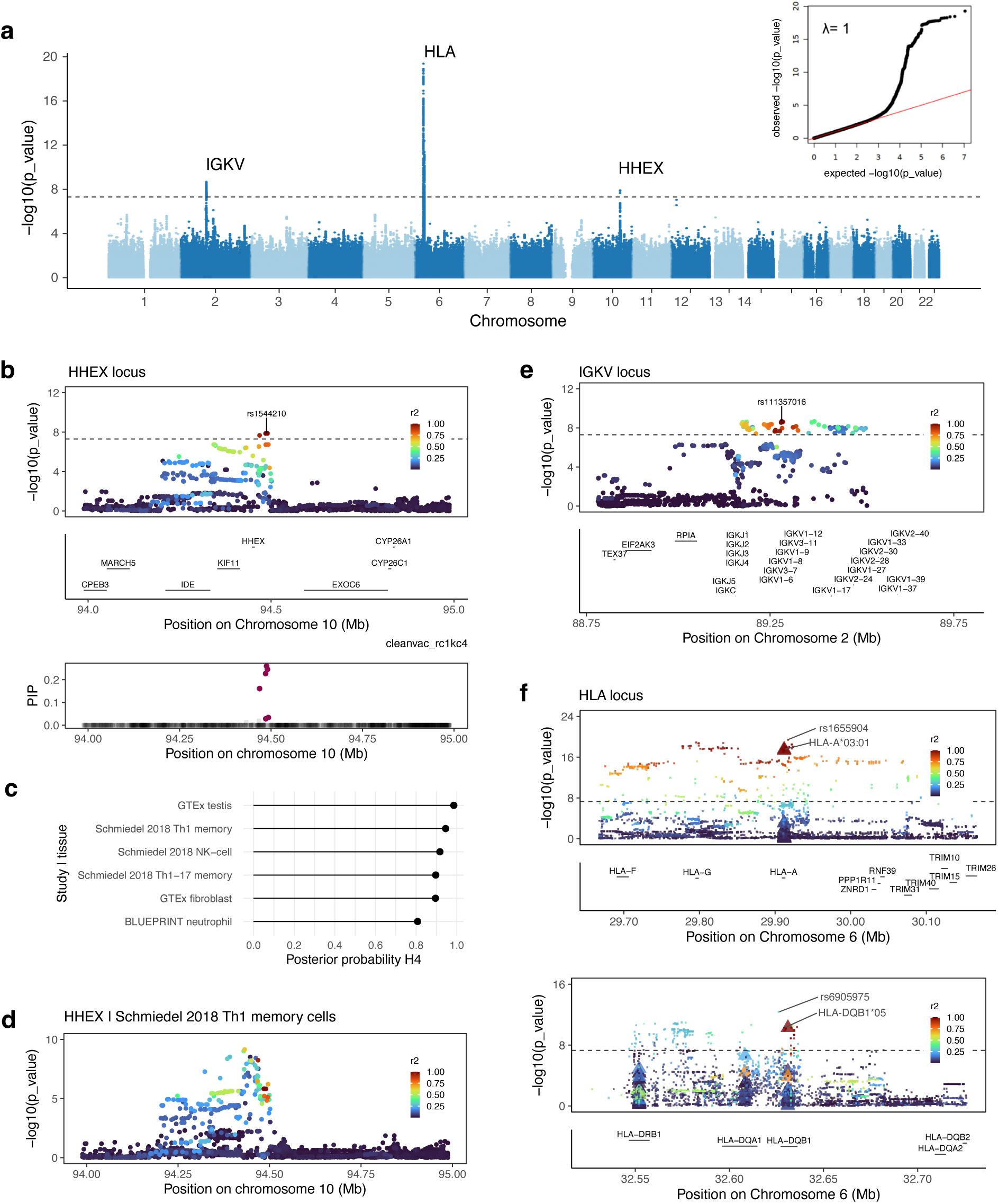
Genetic variation in HLA, IGKV and HHEX loci is associated with differential anti-S antibody concentrations after two COVID-19 vaccinations. **a** Manhattan plot showing the results of the meta-analysis of GWAS for anti-spike antibody concentrations after two COVID-19 vaccinations in infection-naïve individuals across RisCoin and KoCo19 cohorts. Meta-analysis p-value per variant is shown on the y-axis, plotted against the genomic positions of the variants. Genome-wide significant results are annotated with the locus name. The inset shows a qq-plot of observed versus expected association p-values. Inflation factor lambda is noted. **b** Regional association plot (top) of the HHEX locus showing the results of a GWAS meta-analysis of anti-spike antibody concentrations after two COVID-19 vaccinations. Posterior inclusion probability (PIP) values from statistical fine-mapping analysis are shown against the variant position (bottom). Six credible set variants were identified (colored in magenta). **c** Colocalization analyses were performed across all tissues in the eQTL Catalogue. Posterior probability for shared genetic signals (PP4) is shown on the x-axis for six tissues, in which PP4 was higher than 0.8. **d** Regional association plot depicts the eQTL signal for HHEX gene expression measured in Th1 cells. **e** Regional association plot of the IGKV locus showing the results of a GWAS meta-analysis of anti-spike antibody concentrations after two COVID-19 vaccinations. **f** Regional association plot of the HLA class I and II loci depicting the results of a GWAS meta-analysis of anti-spike antibody concentrations after two COVID-19 vaccinations. Analyses were conducted using SNP and HLA allele variation imputed using the T1DGC panel.^36^ SNPs are represented by circles and HLA alleles by triangles. Lead SNP and HLA allele signals per locus are labelled. In panels **b**,**d**,**e**,**f** the color indicates the linkage disequilibrium estimate (r2) to the lead variant or HLA allele and coordinates of the genes in the locus are shown in the lower panel. In all panels, the dashed line represents the genome-wide significance threshold (5×10-8) and variant positions were shown with respect to hg19/GRCH37 genome build.

To prioritise genetic variants and identify effector genes in the novel HHEX locus, we performed statistical fine-mapping and colocalization analysis. Statistical fine mapping identified 1 credible set of 6 variants, with overall low posterior inclusion probabilities, in line with the moderate signal strength of the association analysis (**Fig. 2b**). Colocalization analysis was performed across all tissues in the expression quantitative trait loci (eQTL) Catalogue^33^. We found significant evidence for colocalization, based on posterior probability for shared genetic signals (PP4) larger than 0.8, between the HHEX locus discovered in our study and eQTL signals for the HHEX gene across six different tissues (**Fig. 2c**). Importantly, those included four immune cell types (i.e., Th17, Th1, NK-cells and monocytes), among which the highest PP4 of 0.95 was observed for Th1 cells (**Fig. 2d**). Taken together, these results suggest that the genetic effect of the chromosome 10 locus that we discovered on anti-spike antibody concentrations after two COVID-19 vaccinations is mediated by HHEX gene expression.

Next, we confirmed that the IGKV signal (lead variant rs111357016, GRCh37: chr2:89281528) (**Fig. 2e**) is a replication of the results reported by Sonehara et. al (lead variant rs139861548, GRCh37: chr2:89246954). The lead variant previously reported was not represented in our dataset, so we used a variant in high LD (0.78, 1kg EUR superpopulation^34^) as a proxy for the reported result. Conditioning on either the lead variant in our study or the proxy of the lead variant in Sonehara et. al., we observed a substantial reduction of signal in the whole region (**Supplementary Fig. 2**), indicating that our result is an independent replication of the contribution of the IGKV genetic variation to anti-S antibody level concentrations in a different population. Additionally, we could reproduce the high colocalization of the GWAS signal with eQTL signal for gene IGKV-13 in GTEx^35^ whole blood data (PP4 = 0.94). Taken together, we provided robust evidence for the role of the germline immunoglobulin gene variability in antibody concentrations after two COVID-19 vaccinations and the possible role of immunoglobulin gene expression levels as a mediating factor.

To fine-map the discovered associations in the HLA region, we imputed HLA allele, amino-acid and coding variation using the T1DGC HLA reference panel^36^ and repeated the association analyses for the individual studies.^37^ Meta-analysis of the association runs on the imputed datasets found HLA-A*03:01 as the lead allele signal in the HLA class I region and HLA-DQB1*05 as the lead allele signal in the HLA class II region (**Fig. 2f**). Lead HLA-, amino-acid-and single nucleotide polymorphism (SNP) signals are listed in Table 2. To understand potential functional consequences of variation in amino-acid sequences of the HLA complexes, we located the lead amino-acid signal from our study in a publicly available 3D structure of a HLA-A*02-SARS-CoV-2 peptide complex.^38^ Corroborating other studies^14,17^, we found that the amino-acid, that was the lead signal in the HLA class I region in the association analysis, is located in the peptide binding groove of the HLA complex (**Supplementary Fig. 3**). This suggests that the differential production of spike-specific antibodies in carriers versus non-carriers is due to differential peptide binding affinity and consequently dissimilar CD8+ T-cell activation.

**Table 2.** Summary statistics of the lead HLA allele, amino-acid and SNP variation in the GWAS meta-analysis of anti-spike antibody concentrations after two COVID-19 vaccinations. Genetic effect estimates and association p-values are shown per cohort and for the meta-analysis. Position on chromosome six is given with respect to the hg19/GRCH37 genomic build. Multi allelic variation is encoded as P - presence, A - absence. Biallelic amino-acid variation is encoded with three-letter amino-acid codes. Amino-acid variant names indicate the variable position in the translated protein, and the amino-acid considered in the analysis in case of multiallelic variation. EAF = effect allele frequency.

| Region | Variant | Variant type | Position | Effect allele | EAF | RisCoin beta | KoCo19 beta | Meta-analysis beta | RisCoin p-value | KoCo19 p-value | Meta-analysis p-value |
| --- | --- | --- | --- | --- | --- | --- | --- | --- | --- | --- | --- |
| HLA class I | HLA-A*03:01 | HLA allele | 29911991 | P | 0.15 | 0.30 | 0.22 | 0.26 | 8.0e-13 | 3.1e-07 | 2.7e-18 |
| HLA class I | HLA-A AA 161 | amino-acid | 29911255 | Asp | 0.15 | 0.30 | 0.22 | 0.26 | 2.1e-12 | 2.5e-07 | 5.2e-18 |
| HLA class I | rs1655904 | SNP | 29917146 | C | 0.81 | -0.28 | -0.23 | -0.26 | 4.4e-13 | 1.6e-08 | 4.2e-20 |
| HLA class II | HLA-DQB1*05 | HLA allele | 32631061 | P | 0.18 | -0.19 | -0.14 | -0.17 | 1.0e-08 | 8.3e-04 | 5.3e-11 |
| HLA class II | HLA-DQB1 AA 125 Ser | amino-acid | 32629935 | P | 0.18 | -0.19 | -0.14 | -0.17 | 1.0e-08 | 6.2e-04 | 3.8e-11 |
| HLA class II | rs6905975 | SNP | 32626139 | G | 0.54 | 0.18 | 0.11 | 0.15 | 3.6e-11 | 8.8e-04 | 4.3e-13 |

### Genetic variation in the HLA region associated with altered live-virus neutralization activity against SARS-CoV-2 omicron after two COVID-19 vaccinations

We next assessed if genetic variation was associated with differential antibody-mediated live-virus neutralization activity against SARS-CoV-2 Omicron BA.1 in the sera of individuals who received two vaccine doses in the RisCoin cohort. (**Fig. 3a**). In line with results from previous studies^10,11^, we found a limited predictive value of anti-spike antibody concentrations for live-virus neutralization activities. Indeed, more than a third of participants showed neutralization activity below the threshold of detection (i.e., below a serum dilution of 1:10) while having detectable anti-spike antibody concentrations. In participants, for whom both live-virus neutralization activities and anti-spike antibody concentrations were quantifiable above the assays’ thresholds, the measurements showed a modest yet highly statistically significant correlation (ρ = 0.43, p-value < 1e^-16^) (**Fig. 3b**). These results warranted investigation of the genetical phenotype of the live-virus neutralization activity. Our linear mixed-model association analysis included age, gender, days since vaccination, the first five principal components of genetic data and the genetic relatedness matrix as covariates. The neutralization activity phenotype was inverse-normal transformed. Our GWAS identified two independent, genome-wide significant signals (p-value < 5×10^-8^) on chromosome 6, in the HLA class I and class II regions (**Fig. 3c, d**). Interestingly, discovered lead allele signals, i.e., HLA-A*03:01 and HLA-DQB1*05, were the same as those found in the anti-SARS-CoV-2 spike antibody level analysis (see above). These results suggest that the genetic determinants are shared between the two phenotypes, despite a limited concordance at the phenotype level.

**Fig. 3.**
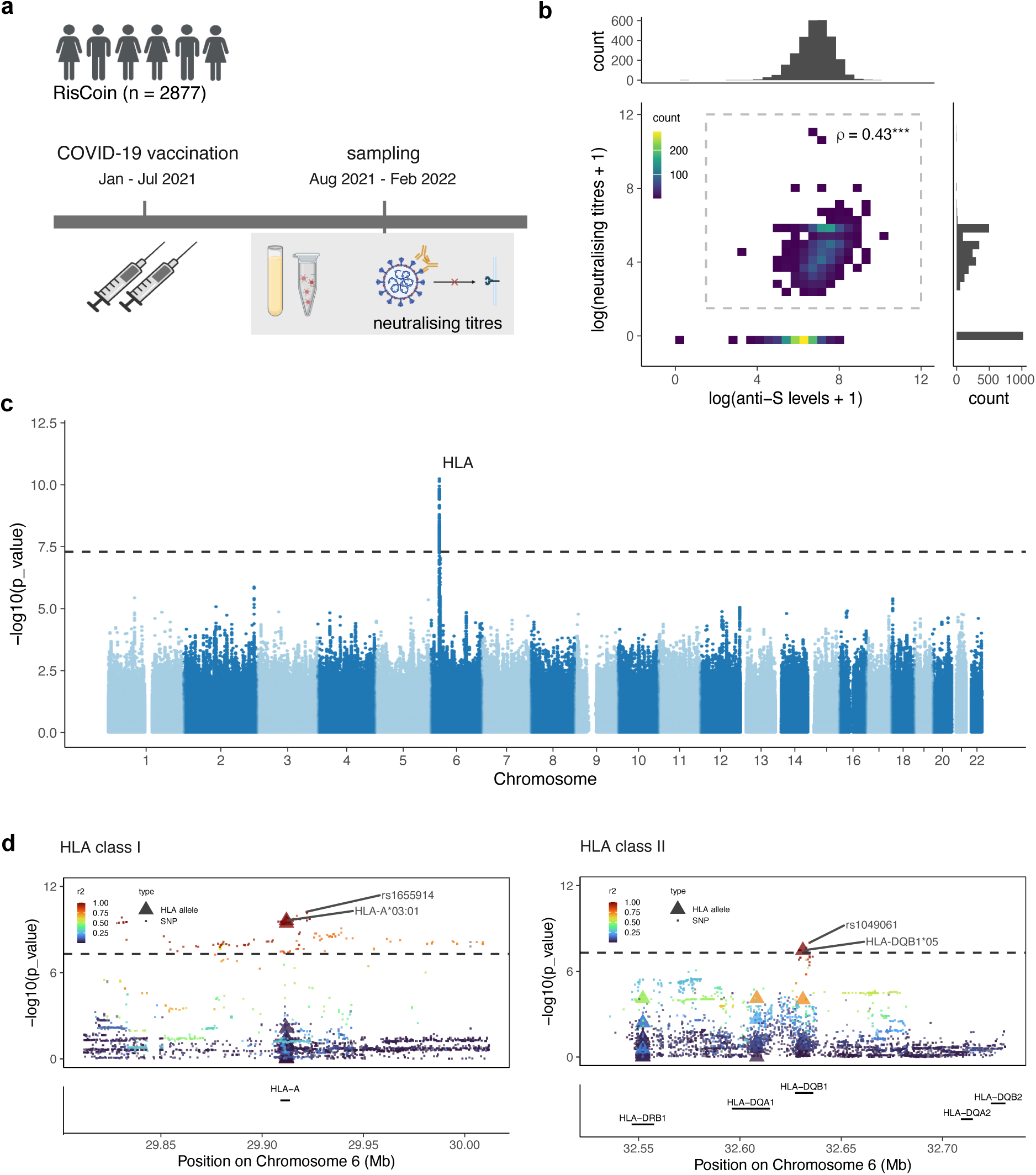
Genetic variation in HLA class I and II regions is associated with differential live-virus neutralization activities after two COVID-19 vaccinations. **a** Cohort overview. We ran GWAS on the live-virus neutralization activity against SARS-CoV-2 Omicron BA.1 from 2877 infection-naïve individuals in the RisCoin cohort, collected at median 227 days post second COVID-19 vaccination. **b** Correlation of anti-spike antibody concentrations and live-virus neutralization activities. **c** Manhattan plot showing the results of the GWAS of live-virus neutralization activity. GWAS p-value per variant is shown on the y-axis, plotted against the positions of the variants on hg19/GRCH37 genomic build. The dashed line represents the genome-wide significance threshold (5×10^-8^). Genome-wide significant results are annotated with the locus name. **d** Regional association plot of the HLA class I (left) and II (right) loci depicting the results of a GWAS conducted using SNP, amino-acid and HLA allele variation imputed using the T1DGC panel.^36^ The color indicates the linkage disequilibrium estimate (r2) to the lead variant. SNPs are represented by circles, and HLA alleles by triangles. Coordinates of the genes in the locus are shown in the lower panel. Lead SNP and HLA allele signals per locus are labelled.

### Genetic variation in HLA class I region associated with differential live-virus neutralization activities and anti-spike antibody concentrations after three COVID-19 vaccinations

We next investigated if genetic contribution to live-virus neutralization and anti-spike antibody concentrations change after receiving a third COVID-19 vaccination. To this end, we ran a GWAS on the RisCoin cohort including data from 1225 individuals who had previously received a third COVID-19 vaccination and were SARS-CoV-2 infection-naïve (**Fig. 4a**). The median time since the third, booster vaccination was 56 days. After the third immunization, the correlation of anti-spike antibody concentrations and neutralization activity against Omicron increased compared to that calculated post-second vaccination (Rho = 0.53) (**Fig. 4b**). Our linear mixed-model association analysis included age, gender, days since vaccination, the first five principal components of genetic data and the genetic relatedness matrix as covariates. Values for anti-spike antibody concentrations and live-virus neutralization activity were inverse-normal transformed. For both phenotypes, we observed one independent genome-wide significant signal (p-value < 5×10^-8^) (**Fig. 4c**), an association in HLA class I region (**Fig. 4d**). The lead allele of this signal, HLA-A*03:01, was identical to that observed for both phenotypes post second vaccination. Of note, the HLA class II signal, HLA-DQB1*05, that we discovered after the second immunization diminished (anti-spike antibody concentrations GWAS p-value = 7×10^-3^) (**Fig. 4e**) albeit showing an effect size (beta = -0.15) comparable to the effect after second vaccination (beta = -0.17). Beyond previously reported associations with post–second vaccination response, our findings demonstrate that the HLA locus influences protective humoral immunity as measured by live-virus neutralization activity and anti-spike antibody concentrations after the third vaccination.

**Fig. 4.**
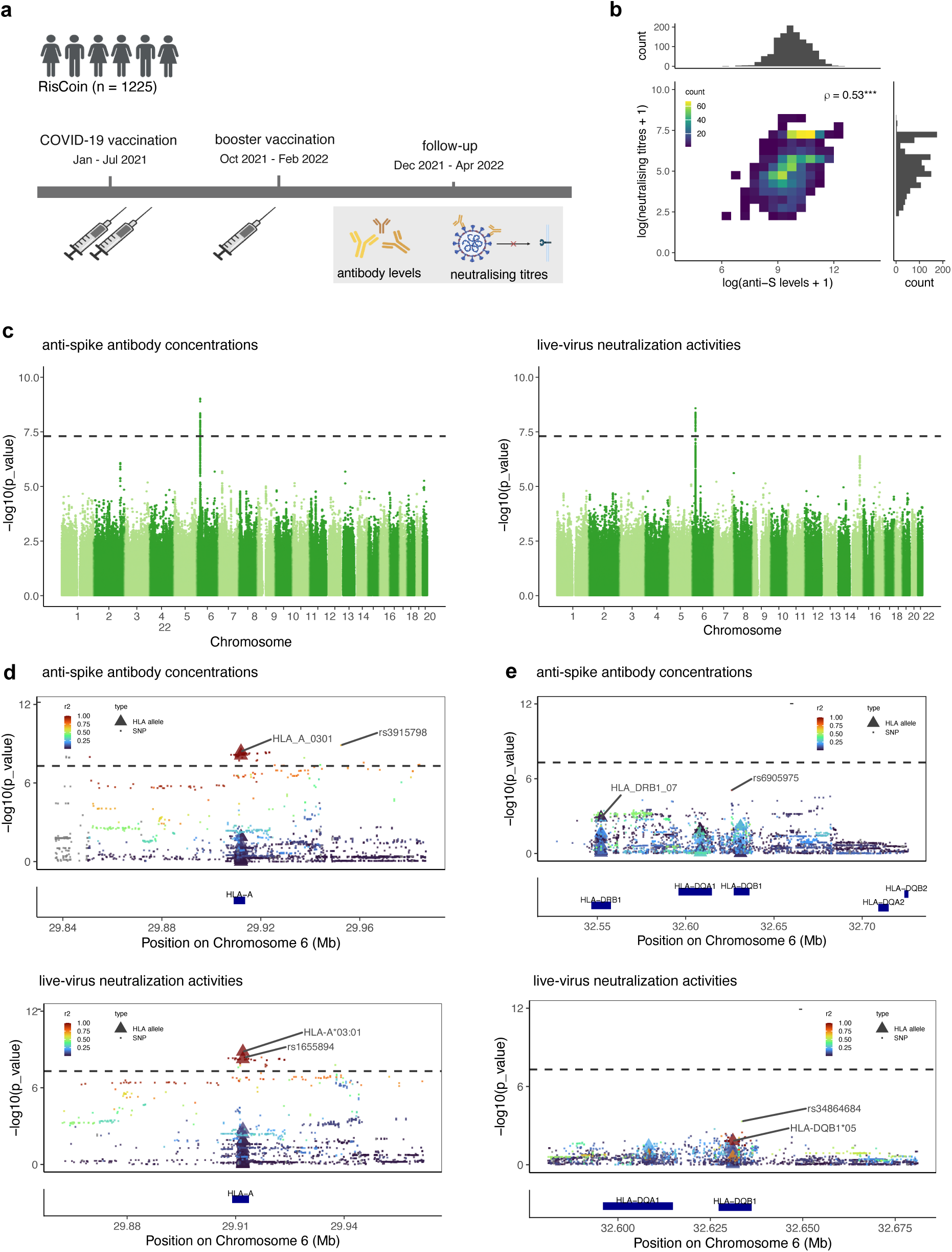
Genetic variation in HLA class I region associates with neutralization activities and anti-spike antibody concentrations after the third COVID-19 vaccination. **a** Cohort overview. We ran GWAS on the live-virus neutralization activity against SARS-CoV-2 Omicron BA.1 and anti-spike antibody concentrations from 1225 infection-naïve individuals in the RisCoin cohort, collected at median 56 days post third COVID-19 vaccination. **b** Correlation of anti-spike antibody concentrations and live-virus neutralization activities. **c** Manhattan plot showing the results of the GWAS of anti-spike antibody concentrations (left) and live-virus neutralization activity (right). GWAS p-value per variant is shown on the y-axis, plotted against the positions of the variants on hg19/GRCH37 genomic build. The dashed line represents the genome-wide significance threshold (5×10^-8^). Genome-wide significant results are annotated with the locus name. Regional association plot of the HLA class I (**d**) and II (**e**) loci, for the anti-spike antibody concentrations (top) and neutralization activities GWAS (bottom), conducted using SNP, amino-acid and HLA allele variation imputed using the T1DGC panel.^36^ The color indicates the linkage disequilibrium estimate (r2) to the lead variant. SNPs are represented by circles, and HLA alleles by triangles. Coordinates of the genes in the locus are shown in the lower panel. Lead SNP and HLA allele signals per locus are labelled.

### Carrying the HLA-A*03:01 allele was associated with decreased incidence of breakthrough infections and increased incidence of self-reported severe vaccination adverse effects

In the RisCoin cohort, self-reported adverse effects of the COVID-19 vaccinations as well as the incidence of SARS-CoV-2 breakthrough infections were assessed utilizing questionnaires and a weekly questionnaire via smartphone app.^39^ We tested whether individual genotypes could predict the incidence of self-reported severe adverse effects or breakthrough infection after receiving three vaccine doses using logistic and Cox proportional hazards regression, respectively. In this analysis, as predictors, we used the genotypes at the lead IGKV and HHEX variants, and lead HLA allele signals identified in the GWAS meta-analysis of anti-spike antibody concentrations (**Table 1. and 2**.). We found that carrying two copies of HLA-A*03:01 was associated with a decreased risk of experiencing a breakthrough infection (hazard ratio = 0.36, 95% confidence interval [0.13, 0.98]), and carrying one copy with an increased risk of experiencing more self-reported severe adverse effects after the third vaccination (odds ratio = 2.22, 95% confidence interval [1.65,2.98]) (**Fig. 5**). These results corroborate previous findings of other studies^40,41^ and demonstrate that beyond neutralizing titres at the third vaccination, the HLA association extends to clinical protection.

**Fig. 5.**
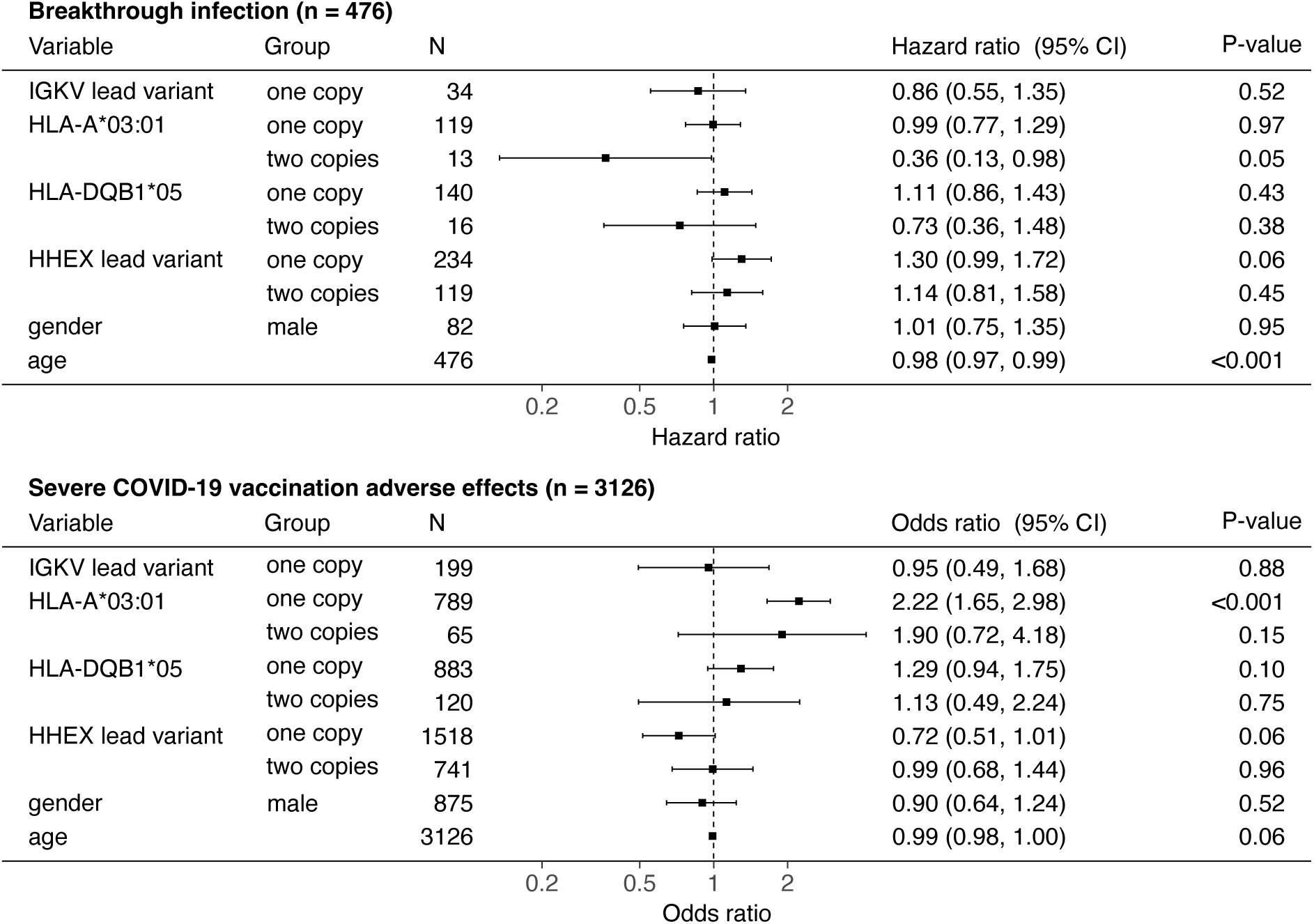
Carrying HLA class I variation is associated with decreased incidence of breakthrough infection and increased incidence of self-reported severe adverse effects after COVID-19 vaccination. Annotated forest plot indicating Cox proportional hazards regression results for breakthrough infection incidence (top) and logistic regression results for self-reported severe adverse effect incidence (bottom) in the RisCoin cohort. From left to right, the plot shows the variable of interest, the group of interest, the number of individuals per variable level, visual and numeric indication of regression results. The results shown are relative to a reference group, defined as individuals with zero copies of the variant of interest, or as females when gender is the variable considered. Boxes indicate hazard ratio estimates for breakthrough infection, and odds ratio estimates for self-reported severe adverse effects incidence. Whiskers indicate the 95% confidence interval. CI = confidence interval.

## Discussion

The genetic contribution to the neutralizing activity of vaccine-induced antibodies and the humoral response after the third vaccination has been unclear. In two large cohorts of predominantly mRNA vaccinees, we show that variants at the HLA, IGKV and HHEX loci influence durable immunity against COVID-19. To our knowledge, this is the first GWAS to quantify neutralizing antibody titres at scale, the largest study to measure anti-spike antibody concentrations, and the first GWAS analysis focused on the third-dose time point in an infection-naïve population. While we replicated and expanded the well-established contribution of HLA, we also identified a novel signal, potentially because of our late sampling time point, underscoring the context specificity of genetic effects on vaccine responses. Our findings at IGKV add to growing evidence that germline immunoglobulin variation shapes vaccine-induced antibody profiles, paralleling historical insights from HLA. Altogether these results may inform design and optimisation of mRNA vaccines beyond COVID-19, including for other infectious diseases and cancer.

The newly discovered association between the HHEX locus and anti-SARS-CoV-2 spike antibody concentrations is, as our findings suggest, likely mediated through differential gene expression levels of the transcription factor HHEX. Of importance, HHEX was recently described as a key player in memory B cell development.^42^ Given that anti-spike specific memory B cells were reported to be good correlates of anti-spike antibody levels several months after COVID-19 vaccination^43,44^, we speculate that HHEX could have an influence on antibody levels via modulation of the memory B cell compartment. Therefore, detection of the HHEX locus may be attributable to our late sampling time point in the RisCoin cohort, i.e. by median 8 months after the second vaccination, when affinity maturation and antibody persistence dominate. It might have also become apparent because we measured continuous antibody levels rather than a binary serostatus. Either the late sampling time point or the continuous measurement, or their combination, could explain why the association between HHEX locus and anti-SARS-CoV-2 spike antibody concentrations was not observed previously and adds to our understanding of long-lasting protection against COVID-19.

Our results regarding the IGKV locus confirm previous reports on the influence of germline variation in the Ig gene on antigen-specific antibody concentrations^17^. Our findings support the hypothesis that the antibody repertoire is constrained by inborn Ig variability. ^45–47^ Individuals carrying specific genetic Ig variants may, thus, be predisposed to produce fewer antibodies after exposure to specific antigens. Narrowing down on the exact genetic variants mediating the effect is currently limited because of the complexity of the IGKV locus, which would require additional sequencing data to resolve. Especially, developing genotype imputation panels for the Ig loci would potentially help overcome this limitation and, thereby, add to the existing evidence for the role of germline Ig variation in antibody repertoire and disease susceptibility.^45–47^

Lead HLA allele signals associated with anti-SARS-CoV-2 antibody responses differ across previously published studies.^14–23^ Population-specific effects have been speculated to play a role in the reported variability, with cross-ancestry meta-analysis being proposed as a way to formally address the issue^48^. In our analyses, HLA-A*03:01 was positively associated with SARS-CoV-2-specific humoral immune responses, whereas HLA-DQB1*05 showed a negative association. In line with previous reports, we suggest that the binding affinity of spike-derived peptides differs between the different HLA-A types. Consequently, HLA-A-dependent CD8+ T-cell activation and downstream immune functions may be affected, ultimately influencing anti-spike antibody production and quality. Interestingly, regarding HLA class II alleles, it was shown *in vitro* that anti-spike memory B cell and overall CD4+ T cell responses differ between HLA-DB1*06 carriers and non-carriers, adding evidence to the proposed mechanism behind the association between HLA variation and antibody concentrations after COVID-19 vaccination.^14^ To our knowledge, we discovered for the first time that HLA loci are not only associated with anti-SARS-CoV-2 spike antibody concentrations after the second COVID-19 vaccination, but also with those after the third shot and, moreover, with live-virus neutralization activities against Omicron BA1. These results highlight the importance of the discovered HLA loci for influencing antibody responses to SARS-CoV-2, even after repeated exposure to the spike antigen, and neutralizing immunity, which was shown to be a strong correlate of protection against symptomatic and severe disease.^6,24,25,49,50^ Participants of our study who were homozygous for HLA-A*03:01 were, indeed, significantly protected from breakthrough infection, in line with reports from others.^40,41^ Collectively, our findings indicate the potential relevance of the HLA locus for future personalised immunization approaches.

Despite elucidating the role of HLA, IGKV and HHEX loci on durable immunity against COVID-19, our study has some limitations. First, the loci highlighted here and elsewhere warrant experimental validation to confirm roles in antibody production and persistence and to assess generalizability across vaccine platforms and antigens. Another limitation is that we only assessed live-virus neutralization activities against Omicron BA.1 and not against other, more recent SARS-CoV-2 variants. There is, however, evidence that vaccination-induced neutralization titres correlate strongly between different virus variants.^10,51^ We, thus, hypothesize that the associations between the HLA loci and live-virus neutralization activity against Omicron BA.1 also apply to other SARS-CoV-2 variants, especially to those of the predominant Omicron clade. Finally, by leveraging a cohort of healthcare workers vaccinated on standardized schedules, we could characterize durability and post–third-dose responses. Nonetheless, power at the third dose was limited with considerably smaller number of participants in the post–third-dose subset compared after the second, and additional loci may emerge with larger sample sizes. Extending genetics to fourth and later boosters would be highly informative as well but would require new data. However, recruiting infection-naïve individuals after ≥3 immunizations became impractical following Omicron, when breakthrough infections were widespread in the general population from Spring 2022 onwards.^52,53^ Consequently, our study might not only be the first but may remain one of very few opportunities to assess genetic determinants of long-lasting immunity against COVID-19.

In conclusion, our study identified novel genetic determinants of humoral immunity to SARS-CoV-2 after the second and third COVID-19 vaccinations. This is the first study to provide relevant context for investigating the genetics of long-lasting antibody responses, neutralizing immunity and humoral immune responses after the third immunization. HLA loci that we showed to be associated with the functional correlate of protection against SARS-CoV-2 but also the other discovered loci may in future be helpful to identify and protect vulnerable individuals and provide them access to personalized vaccination strategies. Some of the genetic variants identified here may not only be important for shaping immunity to SARS-CoV-2 but they may also play a more general role in modulating vaccine-induced immunity to other pathogens.

## Methods

### Study design and participants

The RisCoin cohort comprised 4115 individuals, predominantly healthcare workers, as well as patients diagnosed with inflammatory bowel disease and psychiatric disorders.^28,29^ At enrolment, a questionnaire was filled out and blood samples were donated for serological assessment and genotyping. Healthcare workers who SARS-CoV-2 infection-naive (defined by being negative for anti-nucleocapsid antibodies and self-reporting of never having been tested positive for COVID-19 by PCR), received two doses of mRNA-based COVID-19 vaccine (BNT162b2, mRNA-1273), the second shot at least 28 days ago, took no immunosuppressive medication and had high quality genetic data were selected for genetic analyses at time point #1 (n = 2877). At time point #2, blood samples were taken from participants of the same cohort shortly after the third vaccination. From these, we selected 1225 individuals, who were still SARS-CoV-2 infection-naive and had high quality genetic data, to analyse genetic effects on anti-spike antibody concentrations and live-virus neutralization titres produced after the third immunization (**Fig. 1b**). Weekly questionnaire was conducted using a study app.^39^

The KoCo19 cohort comprised 5313 individuals recruited from randomly selected households in Munich, Germany, for whom anti-SARS-CoV-2 spike and nucleocapsid antibody concentrations and questionnaire data were available at multiple time points throughout the COVID-19 pandemic.^30,31^ We analysed the data from a sampling round that took place between July and October 2021. For genotyping, we picked 1719 participants who had complete data and consent for genetic analyses, received their second vaccination at least 5 days before the sampling round, were SARS-CoV-2 infection-naive as inferred from their anti-nucleocapsid status and donated a dried blood spot sample and of those selected 1654 individuals with high quality genotype data (**Fig. 1b**).

### Antibody detection assays

For blood samples collected in the RisCoin study, serum was extracted by centrifugation at 3,200 x g for 10 min at room temperature. Using the extracted sera we performed the Elecsys Anti-SARS-CoV-2 (Roche, Basel, Switzerland, cat.: 09203095190) and the Elecsys Anti-SARS-CoV-2 S (Roche, cat.: 09289267190) on a Cobas e411 (Roche). Assays were performed in accordance with the manufacturer’s instructions. Following the manufacturer’s recommendations, serum specimens that gave above threshold signals in the Anti-SARS-CoV-2 S assay were diluted in sample buffer and measured again until in range signals were obtained.

For samples collected in the KoCo19 cohort, quantitative anti-spike and anti-nucleocapsid antibody measurements were performed using a self-developed protocol for extraction and measurement in dried blood as described.^54,55^ In brief, participants self-collected dried-blood spot samples (Euroimmun ZV 9701-0101). Antibodies were eluted from the dried-blood spot in 80 µL of PBS buffer containing 5% albumin, 2.5 mM ammonium thiocyanate, and 0.5% Tween20 at 37 °C for 1 hour with shaking at 300rpm. Eluates were analyzed on Cobas e801 units (Roche) using Elecsys Anti-SARS-CoV-2 (Roche) and Elecsys anti-SARS-CoV-2 S (Roche). Dried-blood spot-specific cutoffs and quantitative conversion factors were established and validated against matched plasma samples to ensure accurate serological interpretation.

### Live-virus neutralization assay

As previously described, we used a nasopharyngeal swab to produce high-titer virus stocks of SARS-CoV-2 Omicron BA.1 (GISAID EPI ISL: 7808190).^10,56^ We tested the capacity of serum specimens collected in the RisCoin study to neutralize these virus stocks as described previously.^10,56^ After normalizing the results to the respective abundance of viral RNA in the stocks, a sigmoidal dose-response curve approximation was calculated for each serum tested using Prism 10.2.1 (GraphPad Software, USA) to determine its half-maximal inhibitory concentration (IC50) for live-virus neutralization of SARS-CoV-2 Omicron BA.1. We set the lower threshold for live-virus neutralization to a serum dilution of 1:10.

### Genotype data QC and imputation

Genotype data was obtained using the Infinium Global Screening Array-24 v3.0 (Illumina). We excluded individuals from further analysis with more than 3% missing data, mismatch of self-reported gender and sex information inferred from genetic data, as well as heterozygosity rate of more than 4 standard deviations from the mean heterozygosity rate in the cohort. We then excluded genetic variants with missingness rate greater than 2%, and those deviating from Hardy-Winberg equilibrium (p-value < 1×10-4). Population structure was inspected via principal component analysis and compared to reference samples from the 1kg dataset^34^ (**Supplementary Fig. 3**). In total, 698074 variants and 4068 individuals passed the quality control in RisCoin and 531943 variants and 1850 individuals in the KoCo19 cohort. All quality control metrics were computed with PLINK 1.9.^57^

Genotype imputation was performed on the Munich Imputation Server^58^, where the data was phased with EAGLE2^59^ and imputed with Minimac4^60^ against the Haplotype Reference Consortium (release 1.1) imputation panel.^61^ We selected all variants with imputation scores higher than 0.3 and MAF > 0.05 for further analyses.

### HLA imputation

HLA allele, amino-acid and coding variation imputation was performed using SNP2HLA^62^ against the T1DGC reference panel.^36^ Variants with an imputation score > 0.8 were kept for subsequent analysis.

### Association analysis

Genome-wide association analyses per cohort were conducted in gemma^63^, using a linear mixed model. For each phenotype and time point, we used inverse-normal transformation of the phenotype values. Age, sex, days since most recent vaccination and the first 5 genotype principal components were used as covariates in the model. The relationship between the interval since vaccination and phenotype levels are shown in **Supplementary Fig. 4**. The model also utilized a genetic relatedness matrix to account for relatedness among the participants. For conditional analyses in individual studies, we included the genotype data for the variant that we conditioned for as an additional covariate in the model. HLA association analysis was conducted in gemma, with the same settings as previously used in the genome-wide analysis. We ran the meta-analysis in METAL^64^, using the stderr scheme. We used gcta-cojo^65^ to identify independent associations and perform conditional analyses. Statistical fine-mapping was done using susieR^66,67^. Colocalization was performed with the eQTL Catalogue^33^ gene expression summary statistics using the coloc^68^ package in R.

### Breakthrough infection and self-reported severe adverse effect incidence analyses

The RisCoin cohort participants were invited for serological assessment and to answer a questionnaire several months after the third vaccination. Data from this time point #3 and the weekly app questionnaire were used for evaluating the incidence of breakthrough infections. In the analysis, we included only individuals with exactly three mRNA vaccinations and a known SARS-CoV-2 infection date, in case they experienced a breakthrough infection. Among the 476 individuals with complete data, 310 have experienced a breakthrough infection. Using the data from the initial, follow-up and weekly app questionnaires, we analysed the incidence and severity of participants’ self-reported adverse effects after the third immunization. Among the 3170 individuals with complete data, 207 self-reported severe adverse effects with impairment in everyday life (e.g. doctor’s visit, sick leave). We applied a Cox proportional hazards model using the *coxph* function^69,70^ in R to evaluate the association of breakthrough infection incidence with the four lead variants identified in the anti-spike GWAS meta-analysis. We used logistic regression using the *glm* function^71^ in R to assess the incidence of self-reported severe adverse effects after the third vaccination, with the same lead variants used in the breakthrough infection analysis as predictors. In both models, age and gender were included as covariates.

## Data availability

Single-study GWAS and GWAS meta-analysis summary statistics are currently being deposited in the GWAS Catalog (accession number: tbd).

## Ethics approval and consent to participate

The RisCoin study was approved by the ethics committee of the Faculty of Medicine at the Ludwig Maximilian University of Munich (study-No.: 21–0839). The KoCo19 study protocol was approved by the ethics committee of the Faculty of Medicine at the Ludwig Maximilian University of Munich, Germany (study-No.: 20–275), prior to study initiation. Informed consent was obtained from all study participants prior to study inclusion.

## Author Contributions

Conceptualization and supervision: S.K.-H., S.K. and M.H.; RisCoin study design and funding acquisition: A.C., B.K., H.P.T., K.A., O.T.K., S.K. and V.H.; KoCo19-SEPAN study design and funding acquisition: M.H., E.Z. and C.G.; data generation and curation: P.R.W., C.J., T.G.L.T., R.R.-A., I.-C.K., G.L., M.S., A.Z., N.C., A.W., and J.R.; formal analysis and visualization: B.P.; writing: B.P., P.R.W and S.K.-H.. All authors contributed to editing the manuscript and approved the final version.

## Funding

The RisCoin study was funded by research grants from the German Federal Ministry of Health (Bundesministerium für Gesundheit, BMG) (KA, SK: ZMI1-2521COR933-BMG, the Corona Research Program 21/22 of the Bavarian Ministry of Science and Art (Bayerisches Staatsministerium für Wissenschaft und Kunst) and intramural and extramural funding of participating research groups (AC has been co-funded by the Ministry of Economic and Climate Action #50WB2222, SK has been funded for the KoCo19-CED Study by the Bayerisches Staatsministerium für Wissenschaft und Kunst). The KoCo19 study was funded by the Bavarian State Ministry of Science and the Arts, the University Hospital of Ludwig-Maximilians-University Munich, the Helmholtz Centre Munich, the University of Bonn, the University of Bielefeld, the European Union’s Horizon 2020 research and innovation programme (ORCHESTRA Grant agreement ID: 101016167), Munich Center of Health (McHealth), the Deutsche Forschungsgesellschaft (SEPAN Grant number: HA 7376/3-1), Volkswagenstiftung (E2 Grant number: 99 450) and the German Ministry for Education and Research (MoKoCo19, reference number 01KI20271). Additionally, B. P. is supported by the Helmholtz Association under the joint research school “Munich School for Data Science - MUDS”. BK is the Else Kröner Senior Professor of Pediatrics at University of Munich, financially supported by the charitable Else Kröner-Fresenius-Foundation, LMU Medical Faculty, and LMU University Hospitals. C.G. is supported by the Deutsche Forschungsgemeinschaft, Germany (reference number: GE 2128/3-1). M.H. is supported by Deutsche Forschungsgemeinschaft, Germany (reference number: HO 2228/12-1). S.K.-H. is further supported by the DFG Emmy Noether Programme KI 2091/2-1 (459153572), SFB/TRR237-B29 (369799452), SFB/TRR359-B06 (491676693), BMBF DZKJ (01GL2406A) and an ERC Starting Grant (101076303).

## Competing interests

The authors declare no competing interests.

## Acknowledgements

We thank all members of the RisCoin Study Group (K. Adorjan, O. Keppler, A. Osterman, I. Badell Garcia, M. Huber, P. R. Wratil, A. Gryaznova, T. Jebrini, P. Kohl, S. De Jonge, K. Neumeier, S. Koletzko, B. Koletzko, S. Kim-Helmuth, Y. Hao, J. Horak, S. Koletzko, K. Csollarova, T.G. Le Thi, T. Schwerd, H.P. Török, L. Koletzko, S. Breiteneicher, A. Choukér, M. Tuschen, K. Biere, T. Wöhrle, S. Matzel, M. Hörl, D. Moser, V. Hornung, J. Rech, C. Ludwig, L. Hansbauer, A. Zhelyazkova, M. Klein, S. Völk, S. Kim-Hellmuth, B. Puzek, and G. Kastenmüller). The authors thank all healthcare workers and patients participating in the RisCoin study for their cooperation. We thank the teams of the Emergency Department, LMU University Hospital, LMU Munich (Prof. Dr. M. Klein) and of the Institute for Infectious Diseases and Tropical Medicine, LMU University Hospital, LMU Munich (Prof. Dr. M. Hoelscher, Dr. C. Janke, C. Reinkemeyer, Dr. I. Noreña) for their support in recruiting participants and their collaboration. We thank the leaders of the work package 8 of the COVIM project (Prof. Dr. L. E. Sanders, Berlin and Prof. F. Klein, Cologne) for allowing us to use parts of the questionnaire developed for their study to characterize the participants for later comparison. We are grateful to the board members and colleagues in the administration and medical departments of the University hospital, particularly Prof. Dr. M. Lerch and PD Dr. S. Horster, for providing RisCoin study facilities to recruit hospital employees. We acknowledge the contribution of students, physicians, and scientific staff who helped in the study logistics and recruitment and follow-up of participants (Biener I, Boeing B, Brammer M, Brüseke J, Camci H, Choukér M-T, Choukér M, Csollarova K, D’Amico F, Deutinger M, De Zen F, Faro T, Geist M, Haesner-Stricker C, Han B, Hao Y, Heynckes S, Hölz H, Huppert K, Jurk A, Kaufmann A, Kamm L, Kavrakova I, Knabe R, Klucker E, Kriesel F, Litwin A, Lupoli G, Matzel S, Öztan GN, Rech J, Rosenberger S, Ruf J, Said-Fabry A, Shabani R, Socas K, Späth P, Stern M, Tsvetkova R, Tuschen M, Wohrle T, and Tu L) and medical students. We appreciate Castor EDC for providing us with the electronic data capture system free of charge in their framework of joining the global fight against SARS-CoV-2. We gratefully acknowledge the kind support of our IT expert Wichert S and Dr. Endres S, and the CentraXX project team of KAIROS GmbH in establishing the CentraXX Study App at our LMU University Hospital, LMU Munich. We acknowledge the KoCo19 study team for their contributions to participant recruitment, data collection, and data curation. We acknowledge the members of the Institute of Translational Genomics and the Kim-Hellmuth lab at Helmholtz Munich for their scientific feedback (especially K. Hatzikotoulas, W. Rayner, A. L. Arruda, and E. Yaman) and administrative support (J. Hankinson and A. Weyand). We acknowledge the technical support of Core Facility Genomics at Helmholtz Munich. We thank Peter Lichtner for help with KoCo19 cohort genotyping. We thank N. Fricker for RisCoin cohort genotyping and L. Lin for help with RisCoin genotype data quality control. This research was performed using resources generated by Type 1 Diabetes Genetics Consortium, a collaborative clinical study sponsored by the National Institute of Diabetes and Digestive and Kidney Diseases (NIDDK), National Institute of Allergy and Infectious Diseases (NIAID), National Human Genome Research Institute (NHGRI), National Institute of Child Health and Human Development (NICHD), and Juvenile Diabetes Research Foundation International (JDRF) and supplied by NIDDK Central Repository (NIDDK-CR). This manuscript was not prepared under the auspices of the T1DGC study and does not necessarily reflect the opinions or views of the T1DGC study, study sponsors, NIDDK-CR, or NIH.

## Notes

### Competing Interest Statement

The authors have declared no competing interest.

### Funding Statement

The RisCoin study was funded by research grants from the German Federal Ministry of Health (Bundesministerium fuer Gesundheit, BMG) (KA, SK: ZMI1-2521COR933-BMG, the Corona Research Program 21/22 of the Bavarian Ministry of Science and Art (Bayerisches Staatsministerium fuer Wissenschaft und Kunst) and intramural and extramural funding of participating research groups (AC has been co-funded by the Ministry of Economic and Climate Action #50WB2222, SK has been funded for the KoCo19-CED Study by the Bayerisches Staatsministerium fuer Wissenschaft und Kunst). The KoCo19 study was funded by the Bavarian State Ministry of Science and the Arts, the University Hospital of Ludwig‐Maximilians‐University Munich, the Helmholtz Centre Munich, the University of Bonn, the University of Bielefeld, the European Union's Horizon 2020 research and innovation programme (ORCHESTRA Grant agreement ID: 101016167), Munich Center of Health (McHealth), the Deutsche Forschungsgesellschaft (SEPAN Grant number: HA 7376/3‐1), Volkswagenstiftung (E2 Grant number: 99 450) and the German Ministry for Education and Research (MoKoCo19, reference number 01KI20271). Additionally, B. P. is supported by the Helmholtz Association under the joint research school "Munich School for Data Science - MUDS". BK is the Else Kroener Senior Professor of Pediatrics at University of Munich, financially supported by the charitable Else Kroener-Fresenius-Foundation, LMU Medical Faculty, and LMU University Hospitals. C.G. is supported by the Deutsche Forschungsgemeinschaft, Germany (reference number: GE 2128/3-1). M.H. is supported by Deutsche Forschungsgemeinschaft, Germany (reference number: HO 2228/12-1). S.K.-H. is further supported by the DFG Emmy Noether Programme KI 2091/2-1 (459153572), SFB/TRR237-B29 (369799452), and an ERC Starting Grant (101076303).

### Author Declarations

Ethics committee of the Faculty of Medicine at the Ludwig Maximilian University of Munich, Germany gave ethical approval for this work.

