## Supplementary Material for "Genetic variation in HLA, IGKV and HHEX loci influence mRNA vaccine-induced long-lasting humoral protection against COVID-19"

**Suppl. Table 1 | Characteristics of RisCoin and KoCo19 individuals selected for the genetic analyses of humoral response after two (Timepoint 1) or three (Timepoint 2) COVID-19 vaccinations.** For KoCo19, anti-spike concentrations were converted to BAU using the formula from Castelletti et. al[1]

| **Characteristic** | **RisCoin Timepoint 1 Post second vaccination** N = 2,877[1] | **KoCo19 Timepoint 1 Post second vaccination** N = 1,654[1] | **RisCoin Timepoint 2 Post third vaccination** N = 1,225[1] |
|---|---|---|---|
| age | 39 (30, 52) | 52 (38, 63) | 44 (33, 55) |
| gender | | | |
| female | 2,141 (74%) | 888 (54%) | 943 (77%) |
| male | 736 (26%) | 766 (46%) | 282 (23%) |
| days since last vaccination | 227 (187, 263) | 46 (27, 66) | 56 (49, 63) |
| log(anti-S concentration) | 6.87 (6.27, 7.40) | 8.23 (7.45, 8.81) | 9.73 (9.17, 10.38) |
| log(live virus neutralization activity) | 3.91 (0.00, 5.00) | NA (NA, NA) | 5.17 (4.36, 6.14) |
| **vaccine 1** | | | |
| Comirnaty (BioNTech/Pfizer) | 2,877 (100%) | 1,114 (67%) | 1,225 (100%) |
| COVID-19 Vaccine (Moderna) | 0 (0%) | 144 (8.7%) | 0 (0%) |
| Vaxzevria (AstraZeneca AB) | 0 (0%) | 396 (24%) | 0 (0%) |
| Others | 0 (0%) | 0 (0%) | 0 (0%) |
| **vaccine 2** | | | |
| Comirnaty (BioNTech/Pfizer) | 2,877 (100%) | 1,295 (78%) | 1,225 (100%) |
| COVID-19 Vaccine (Moderna) | 0 (0%) | 220 (13%) | 0 (0%) |
| Vaxzevria (AstraZeneca AB) | 0 (0%) | 139 (8.4%) | 0 (0%) |
| Others | 0 (0%) | 0 (0%) | 0 (0%) |
| **vaccine 3** | | | |
| Comirnaty (BioNTech/Pfizer) | 0 (0%) | 0 (0%) | 816 (67%) |
| COVID-19 Vaccine (Moderna) | 0 (0%) | 0 (0%) | 409 (33%) |
| not applicable | 2,877 (100%) | 1,654 (100%) | 0 (0%) |
| [1]Median (Q1, Q3); n (%) | | | |

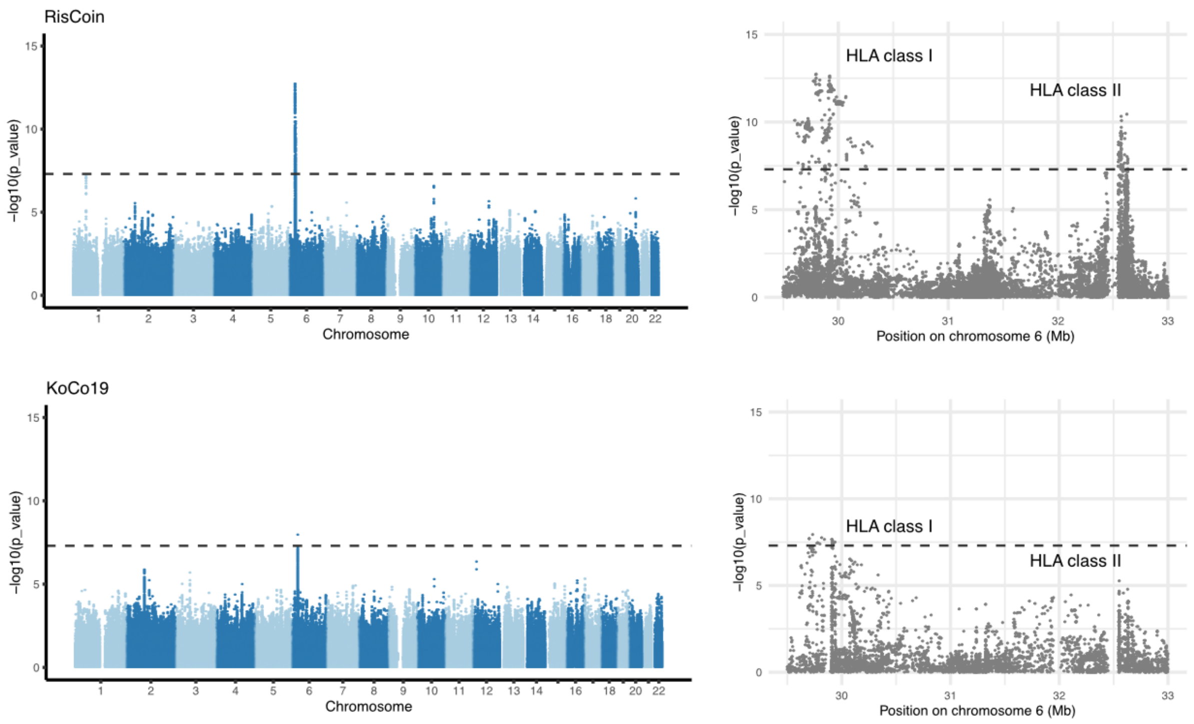


**Suppl. Fig. 1 | Single-study GWAS results.** Manhattan plot (left) and HLA regional association plot (right) showing the results of the single-study GWAS for anti-spike antibody concentrations after two COVID-19 vaccinations in infection-naïve individuals in RisCoin (top) and KoCo19 (bottom) cohorts. GWAS p-value per variant is shown on the y-axis, plotted against the positions of the variants on hg19/GRCH37 genomic build. The dashed line represents the genome-wide significance threshold ($5x10^{-8}$). HLA class I and II regions are indicated with a label.

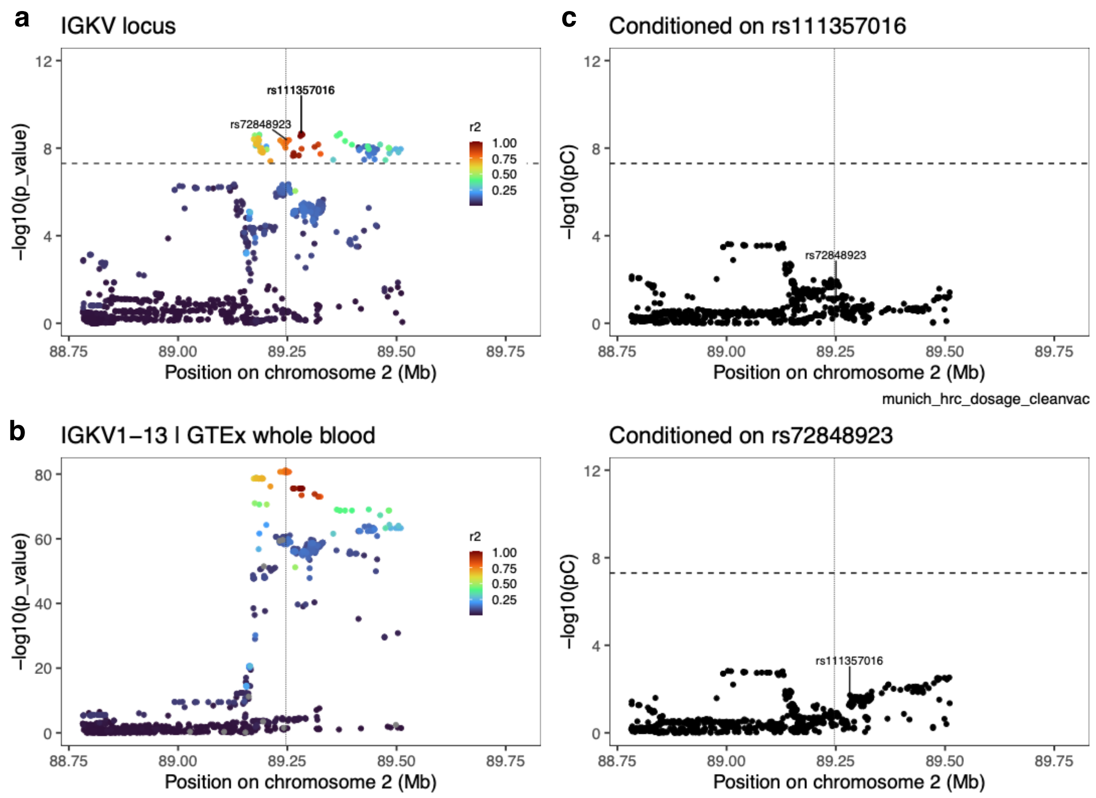


**Suppl. Fig. 2 | Chromosome 2 IGKV locus plot. a** Regional association plot of the IGKV locus showing the results of a GWAS meta-analysis of anti-spike antibody concentrations after two COVID-19 vaccinations. Meta-analysis p-values per variant are shown on the y-axis against the positions of the variants on hg19/GRCH37 genome build. The dashed line represents the genome-wide significance threshold ($5x10^{-8}$). The color indicates the linkage disequilibrium estimate (r2) to the lead variant. **b** Regional association plot of the IGKV locus for IGKV1-13 gene expression QTL data from GTEx. **c** Regional association plot of the IGKV locus showing the results of conditional analysis, conditioning on the lead variant from our study (top) and a proxy of lead variant from Sonehara et. al.[2] (bottom)

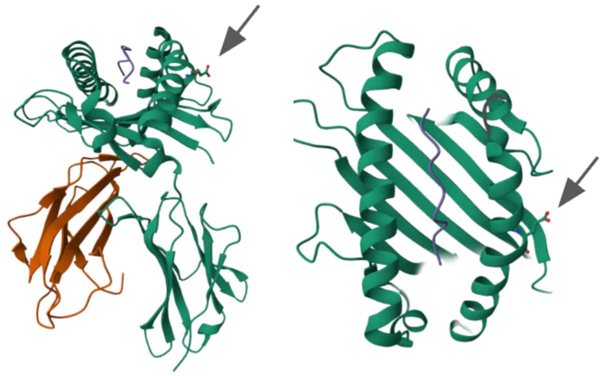

**Suppl. Fig. 3 | Crystal structure of the HLA-A*02 and SARS-CoV-2 peptide complex.** A side- (left) and top- (right) view of the 3D crystal structure of HLA-A*02, in green, presenting a SARS-CoV-2 spike derived peptide, in purple.[3] The position of the lead amino-acid signal from the genetic analysis (HLA-A position 161 D>E) is indicated with an arrow. Of note, the amino-acid position 161 of HLA-A*02 and *03 are identical.

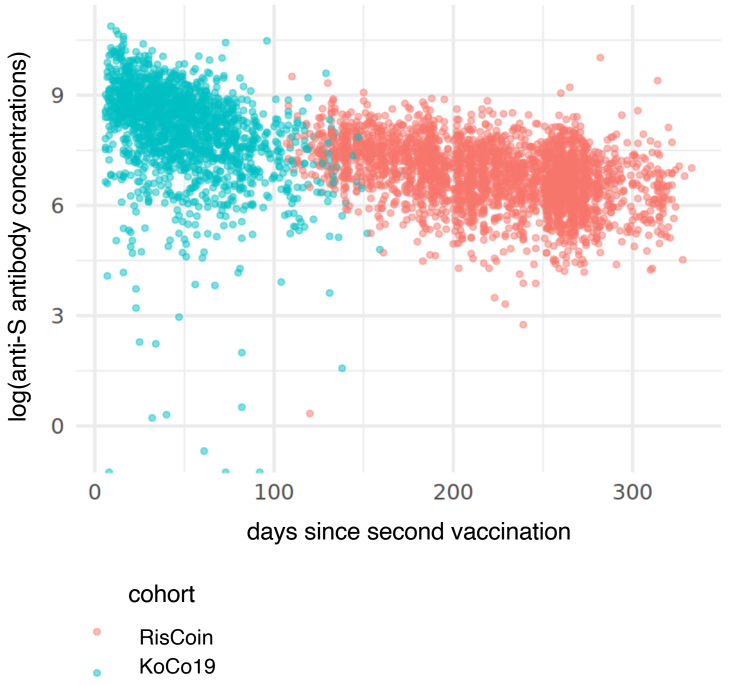


**Suppl. Fig. 4 | Anti-spike antibody concentrations versus time since last vaccination for the RisCoin and KoCo19 cohorts.** For KoCo19, DBS anti-spike values were converted to BAU using the formula from Castelletti et. al[1]

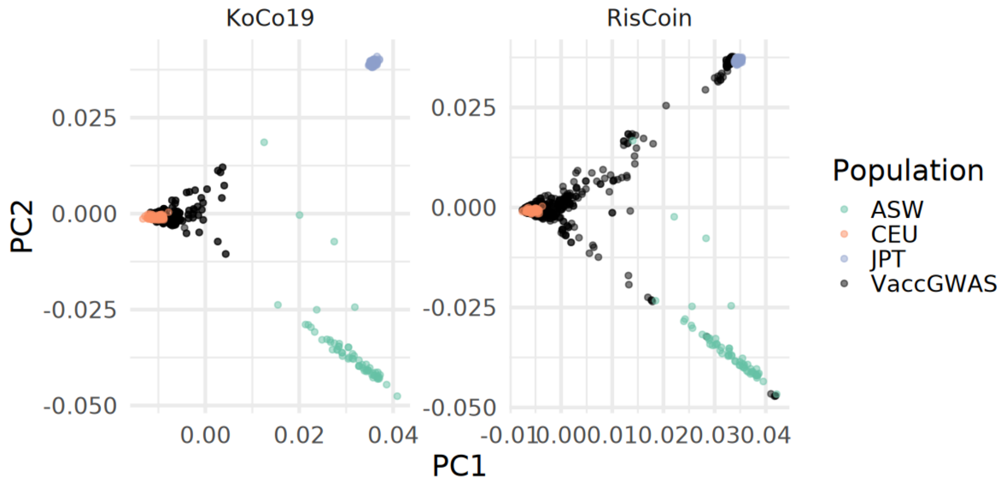


**Suppl. Fig 5 | PCA of genotype data for KoCo19, RisCoin and reference populations from the 1kg dataset.** Two first principal components of genotype data of individuals from the RisCoin, Koco19 cohorts and the 1kg[4] reference dataset are shown on the x and y axis. The color of the points indicates the population.
